# Protocol for a Feasibility study incorporating a Randomised Pilot Trial with an Embedded Process Evaluation and Feasibility Economic Analysis of ThinkCancer!: A primary care intervention to expedite cancer diagnosis in Wales

**DOI:** 10.1101/2020.12.01.20241554

**Authors:** Stefanie Disbeschl, Alun Surgey, Jessica L Roberts, Annie Hendry, Ruth Lewis, Nia Goulden, Zoe Hoare, Nefyn Williams, Bethany Fern Anthony, Rhiannon Tudor Edwards, Rebecca-Jane Law, Julia Hiscock, Andrew Carson-Stevens, Richard D Neal, Clare Wilkinson

## Abstract

**Background:** Relative to the rest of Europe, the UK has relatively poor cancer outcomes, with late diagnosis and a slow referral process being major contributors. General practitioners (GPs) are often faced with patients presenting with a multitude of non-specific symptoms that could be cancer. Safety netting can be used to manage diagnostic uncertainty by ensuring patients with vague symptoms are appropriately monitored, which is now even more crucial due to the ongoing Covid-19 pandemic and its major impact on cancer referrals. The ThinkCancer! Workshop is an educational behaviour change intervention aimed at the whole general practice team, designed to improve primary care approaches to ensure timely diagnosis of cancer. The workshop will consist of teaching and awareness sessions, the appointment of a Safety Netting Champion and the development of a bespoke Safety Netting Plan, and has been adapted so it can be delivered remotely. This study aims to assess the feasibility of the ThinkCancer! Intervention for a future definitive randomised controlled trial.

**Methods:** The ThinkCancer! study is a randomised, multisite feasibility trial, with an embedded process evaluation and feasibility economic analysis. Twenty-three to 30 general practices will be recruited across Wales, randomised in a ratio of 2:1 of intervention versus control who will follow usual care. The workshop will be delivered by a GP educator and will be adapted iteratively throughout the trial period. Baseline practice characteristics will be collected via questionnaire. We will also collect Primary Care Intervals (PCI), Two Week Wait (2WW) referral rates, conversion rates and detection rates at baseline and six months post-randomisation. Participant feedback, researcher reflections and economic costings will be collected following each workshop. A process evaluation will assess implementation using an adapted Normalisation Measure Development (NoMAD) questionnaire and qualitative interviews. An economic feasibility analysis will inform a future economic evaluation.

**Discussion:** This study will allow us to test and further develop a novel evidenced-based complex intervention aimed at general practice teams to expedite the diagnosis of cancer in primary care. The results from this study will inform the future design of a full-scale definitive phase III trial.

**Trial registratio:** intended registry: clinicaltrials.gov

## INTRODUCTION

### Background

Cancer survival in the UK lags behind other western countries.^1^ Referral rates and adherence to guidelines are lower,^2,3^ primary care providers (PCPs) are less likely to take action on potential cancer symptoms^4^ and cancer tends to be diagnosed at a later stage, often only after patients have presented to acute or emergency secondary care services.^3^ Compared with the rest of Europe, the UK has relatively low one-year survival, which could be due to later diagnosis.^5^

Timely diagnosis is key to improving cancer outcomes^6^ and cancer survival.^7^ Earlier diagnosis could also reduce the proportion of cancer patients diagnosed through emergency care.^7^ Early diagnosis is a rising priority in cancer policy,^6,8^ because it is cost-effective^1^ and the incidence of cancer is increasing.^9^ Policies in Wales emphasise the importance of early diagnosis, and recognise that increasing demand and a slow referral process are significant barriers to a quick cancer diagnosis.^3^ Following the implementation of these policies, progress has been slow, with late stage diagnosis continuing to be an issue.^10^

The timely diagnosis of cancer has become even more crucial as we enter a period in which primary care and cancer management has changed dramatically due to the ongoing Covid-19 pandemic.^11-13^ Early figures have shown a 76% decrease in urgent cancer referrals across the UK and predict a 20% increase in excess deaths for patients with newly diagnosed cancers.^14^ There has also been a drop in the number of patients presenting to primary care^15^; patients and clinicians may have concerns due to a perceived risk of contracting Covid-19 in a healthcare setting.^14^ Some patients may also attribute symptoms that could be cancer to Covid-19 and therefore, may avoid health services altogether.^11,13,16^ General practitioners (GPs) may also be reluctant to refer patients on to secondary care in order to minimise infection risk.^15^ The increased use of remote consultation as a result of the pandemic will also have implications for the early diagnosis of cancer, as important consultation techniques such as the use of visual cues and physical examination may be impacted. These issues highlight the ever-important need of safety netting, and Covid-related considerations will need to be made by GPs in their safety netting approaches.^11^

Primary care providers (PCPs) play a vital role in the early diagnosis of cancer.^17^ A key diagnostic stage is the Primary Care Interval (PCI), which is the time from first presentation to a GP with a symptom that could be cancer, to the subsequent referral to a specialist in secondary care.^18^ However, with an ever expanding role, PCPs are presented with a plethora of non-specific symptoms, of which only a small proportion are caused by cancer, and many overlap with other diseases.^6^ Furthermore, with certain cancers, patients may not present with any alarm symptoms^6,17^ which often results in a delayed cancer diagnosis.^2^ In addition, guidelines to expedite early cancer diagnosis are often unclear, with great variation in strategies between different GPs.^19^

### Rationale and previous work

Clinical behaviour change interventions targeting PCPs have the potential to address barriers to suspected cancer identification and referral, and could expedite the diagnosis of cancer and improve cancer outcomes overall.^20^ There is some evidence that educational interventions targeted at PCPs could reduce the PCI, providing they encompass a multidimensional, interactive and tailored approach.^20-22^ A recent systematic review of primary care interventions suggested that a whole-practice approach providing opportunities for peer review and feedback could have a positive effect on referral practices, in addition to existing guidelines being revisited through training and reinforcement.^23^

While the timely diagnosis of cancer is crucial, urgent referral can lead to over-diagnosis and over-investigation which can be harmful to the patient.^7,24^ This risk is especially high if the patient presents with vague symptoms.^25^ Safety netting, a tool used to manage diagnostic uncertainty,^26^ can address these issues by ensuring that patients with non-specific symptoms are not ignored.^24^ Instead of immediate referral, patients are monitored according to a set step-wise investigational plan, while ensuring they are referred in a timely manner as and when required.^24^ Although safety netting is currently recommended by national guidelines,^27^ there are no clear recommendations on how to do it.^19,24,26^

In summary, it is relevant and befitting to develop and test interventions aimed at improving the quality and consistency of primary care approaches to ensure timely diagnosis of cancer in the UK. This will require multicomponent and complex behavioural change interventions, which utilise a multidimensional, interactive, tailored, whole-practice approach.

The ThinkCancer! intervention is a complex behaviour change intervention aimed at general medical practice teams, developed as part of the Wales Interventions for Cancer Knowledge and Early Diagnosis (WICKED) research programme, described in more detail elsewhere.^28^ It consists of an educational workshop that includes early diagnosis and awareness sessions, evaluation of current practice-based safety netting systems and the appointment of a safety netting champion. The workshop will be led by an educational facilitator who will guide the development of a bespoke safety netting plan for each practice.

The aim of this study is to assess the feasibility of delivering the ThinkCancer! intervention and conducting a future, definitive randomised controlled trial (RCT) to assess effectiveness and cost-effectiveness, in order to establish whether the intervention can be rolled out in practice.

## STUDY OBJECTIVES

The objectives of this study are as follows:

1. To assess the feasibility of a future definitive RCT by monitoring recruitment and retention, outcome measure completion and reasons for decline.
2. To assess the acceptability, feasibility, and utility of the ThinkCancer! intervention as a whole, and of each of its individual components, and refining the intervention as necessary.
3. To determine the most appropriate primary outcome measure for a definitive RCT and producing means and confidence intervals for calculating effect sizes for the design of a definitive trial.
4. To describe current contextual differences, and similarities, between general medical practices and their usual safety netting practices.
5. To identify and test the methods and outcome measures for a process evaluation of a future definitive RCT.
6. To undertake a feasibility analysis of the most appropriate approach for an economic evaluation alongside a future definitive trial.

## METHODS

### Study design

The feasibility study incorporates a pragmatic, multisite, two-armed, superiority, pilot RCT. There is an embedded process evaluation and feasibility economic analysis. The unit of randomisation is the general medical practice, and the primary clinical outcome is collected at the practice level.

The term ‘feasibility’ is used in accordance with the conceptual framework developed by Eldridge and colleagues,^29^ where it is described as an umbrella term within which pilot trials are a component. Furthermore, the study has been designed in accordance with the MRC Framework for evaluating complex interventions.^30^ The trial will be conducted according to NIHR guidance,^31^ and recommendations for good practice in pilot studies.^32^ The process evaluation, which will be based on a mixed-methods approach, will follow the MRC guidance for process evaluations of complex interventions.^33^ During the initial piloting or feasibility testing stage of an intervention, process evaluation has a vital role in understanding and planning the future potential implementation of the intervention and optimising its design and evaluation.^33^

The feasibility economic analysis will explore the appropriate future perspective of analysis; most appropriate methods of gathering costs; range and value of outcome measures, and undertake a feasibility budget impact analysis of the ThinkCancer! intervention developed through a range of blended methods that it is delivered online (either in a live format or pre-recorded) or face-to-face in general practices across north Wales.

This study protocol was developed in line with the Standard Protocol Items: Recommendations for Interventional Trials (SPIRIT) guidelines^34^; the SPIRIT checklist (appendix 1) and the schedule of procedures can be seen in Table1. The SPIRIT checklist has been adapted in accordance with the CONSORT extension to pilot and feasibility trials.^35^

**Table 1:**
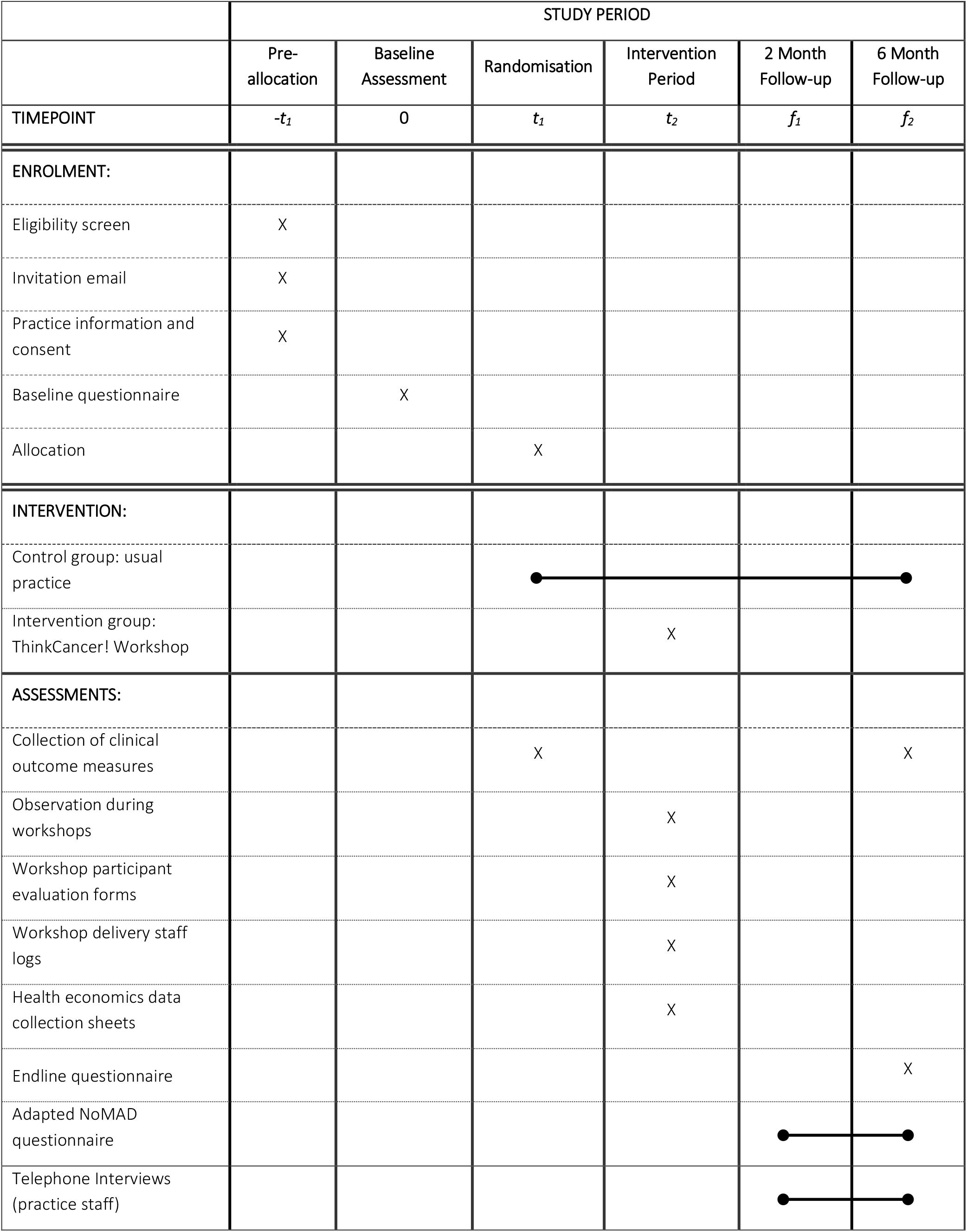
SPIRIT protocol schedule of procedures for the ThinkCancer! study

### Study setting

The setting for this study is primary care. The intervention will be delivered in-practice or online to individual general medical practices and incorporates a whole-practice approach. The trial will be conducted across Wales and practices will be recruited from all seven Welsh health boards.

### Intervention

The proposed intervention, the ThinkCancer! Workshop, has four chief components. The first are two educational sessions, one for all clinical staff (the ‘early diagnosis’ session) and one for non-clinical but patient-facing staff (the ‘cancer aware’ session). The early diagnosis session is delivered as a teaching seminar with learning outcomes focussed on NICE NG12 Suspected Cancer: recognition and referral guidelines^27^, hot topics exploring the harder to recognise cancer presentations and consultation-level safety netting. As a proposed aid to support and formalise safety netting, a new tool – the Symptom Safety Netting Action Plan (SSNAP) will be introduced. This session will also see the introduction of the ThinkCancer! Handbook, which will contain all the resources used in the workshop as well as external resources regarding early diagnosis and safety netting, such as NICE guidance and online learning resources. The cancer aware session is less formal with more convenor-led discussion around cancer red flag symptoms that non-clinical staff may encounter. The secondary aim of this session is to gauge and explore issues and norms around raising concerns within the practice team. The third session (the ‘safety netting session’) involves the two final components of the intervention, the co-production of a bespoke Cancer Safety Netting Plan (CSNP) and appointment of a Cancer Safety Netting Champion (CSNC). This session is attended by a combination of clinical and administrative staff who will be involved in the design and implementation of a new plan.

Members of the research team will deliver the intervention; the GP Educator (AS) will oversee the workshop, supported by up to two researchers. The workshop was originally designed to be delivered face-to-face in participating practices during practices’ allocated protected time for educational and professional development. However, due to the ongoing Covid-19 pandemic, the workshop has been adapted into a digital format and can

be delivered in one of three ways: (i) fully remote and live via online conferencing platforms; (ii) blended delivery, where practices are offered a combination of pre-recorded versions and live remote delivery of the different sessions; and (iii) face-to-face in the practice, as originally intended, if the situation allows. Practices that opt for the blended delivery of the workshop can choose to receive pre-recorded videos of sessions 1 and 2, presented by the GP Educator, allowing participants to engage with the materials in their own time. The final session, which focuses on the Cancer Safety Netting Plan, will still need to be delivered as a live session due to its interactive components. There will be flexibility to work with the practice to allow the bespoke design of the workshop format to improve the reach of the intervention. Practices receiving the intervention in any of the remote forms will be sent all of the workshop materials, including the handbook and SSNAP tool, via post. If there is the possibility to deliver the workshop face-to-face, materials will be distributed in the practice at the beginning of the workshop. Practices randomised to the control group may also receive the pre-recorded videos at the end of the study period, along with the intervention materials.

A logic model (figure 1) has been developed to describe the intervention components and how they link to the intended outcomes and will be adapted throughout the study period.

**Figure 1:** ThinkCancer! Logic model

### Outcome Measures

The outcomes that will be reported in this feasibility study are as follows:

Recruitment will be assessed quantitatively by capturing the numbers of practices

▪ approached,
▪ interested in participating,
▪ consented,
▪ randomised

A description of the excluded practices will be included to help identify potential future eligibility criteria and reasons for non-participation will be sought.

Retention will be assessed quantitatively by the numbers of practices providing both baseline and follow up data. Data on individual practice characteristics will be collected to:

▪ describe the studied sample,
▪ identify potential effect modifiers,
▪ allow identification of ‘usual practice’

The overarching goal of the ThinkCancer! Intervention is to change GPs’ and primary care practices’ behaviours and systems, encouraging them to be more cancer-aware and act sooner on clinical presentations that could be cancer, or more effectively safety net cases where concern exists but criteria for immediate referral are not met. The proposed primary clinical outcomes for the definitive RCT relate to the early referral of suspected cancer. They include the two-week wait (2WW) referral rate and the PCI. The 2WW referral rate is defined as the crude rate of 2WW referrals multiplied by 100,000 and divided by practice list size.^36^ The PCI is defined as the time between the date of first presentation and the date of referral.^18^

Secondary clinical outcome measures include the conversion rate and the detection rate. The conversion rate is defined as the “proportion of 2WW referrals that are subsequently diagnosed with cancer”^36^ and the detection rate consists of the “proportion of new cancer cases treated who were referred through the Two Week Wait route”,^36^ also known as the sensitivity.^37^ These measures will allow us to further explore potential clinical outcomes.

Although it is unlikely that we will detect a difference in the clinical measures in the feasibility study due to the small sample and short duration of follow-up, we do expect these outcome measures to be affected by the ongoing Covid-19 pandemic. However, the feasibility of using these outcomes will be assessed.

Feasibility of using potential clinical primary outcomes, 2WW referral rate, PCI, conversion rates and detection rates, will be assessed quantitatively by determining:

▪ ability to collect/capture data from the practices, based on completion rates of data collection forms
▪ ability to extract relevant data from routinely collected data at Health Board level and from individual practices, based on whether we are able to obtain the data via Health Board contacts and whether these data are comparable with those collected by individual practices
▪ suitability and variability of the data to perform as primary outcome measures for clinical effectiveness

Acceptance, adherence to and fidelity of the intervention will be assessed by:

▪ reviewing participant views, reflections and perceptions expressed via post-workshop feedback forms and interviews
▪ post-workshop reflections from the intervention delivery staff
▪ the ability to organise/schedule and deliver workshops
▪ the number of practices that actively nominate a safety netting champion
▪ the number of practices that demonstrate the use of the safety netting plan

To inform the process evaluation for the future definitive trial, we will:

▪ evaluate how to scale up for any future process evaluation,
▪ determine the acceptability and appropriateness of the interview process
▪ identify barriers and facilitators to successful implementation,
▪ test measures for assessing reach, dose and fidelity of the intervention

For the health economics component we will:

▪ determine the feasibility of collecting data relating to the costing of the intervention via costings forms
▪ identify variables necessary for the design of a future economic evaluation alongside a definitive trial
▪ review relevant literature on the cost-effectiveness of online and mixed-methods Continuing Professional Development (CPD) programmes for health professionals in a community setting
▪ consult the DIRUM database^38^ to identify the most appropriate way of capturing the costs of the ThinkCancer! Intervention (online delivery and mixed-methods delivery).

Adaptations to the intervention and logic model will also be recorded throughout the trial period.

#### Progression criteria

The outcome measures relating to recruitment, retention and adherence/fidelity will be assessed using RAG criteria, as defined below:

1. Confirmation of adequate recruitment for a definitive trial at practice level. Go: 20 or more general medical practices recruited; Review: 15-19 recruited; Stop: < 15 practices recruited.
2. Confirmation of adequate retention for the definitive trial at practice level. Go: 80% or more practices retained; Review: 65-79% practices retained; Stop: < 65% practices retained
3. Confirmation of adequate fidelity of the intervention. Go: 80% or more of all intervention sessions delivered, Review: 50-79% of all interventions delivered; Stop: < 50% of interventions delivered.
4. Confirmation of adequate fidelity at individual practice staff level: From each general practice:
  a. at least 50% of the clinical staff should attend the workshops;
  b. at least 50% of the administrative staff should attend the workshops, comprising at least 50% of the reception and secretarial staff as well as the practice manager.
  c. Staff who do not attend the training should have the information cascaded to them by a member of the team who did attend the training. At least 75% of the staff should receive the training either directly or indirectly.

Progression criteria relating to obtaining data regarding completion of outcome measures will be assessed using the following progression criteria:

1. Routine data. Go: data from 70% or more practices obtained; Review: data from < 70% of practices obtained
2. Individual data. Go: data from 70% or more of individuals of each practice obtained; Review: data from < 70% of individuals from each practice obtained

These criteria would reflect the potential outcomes for exclusion/inclusion at a full definitive trial stage rather than prevention of the study progressing.

### Sample size

We aim to recruit 23-30 general medical practices, depending on the ease of recruitment, using a randomisation allocation ratio of 2:1 of intervention versus control. Randomising in preference to the intervention will allow us to iteratively develop the intervention more effectively.

As this is a feasibility study, there is no requirement for a formal power calculation. This study is not intended to be powered to identify a clinically meaningful difference between the intervention groups for the primary outcome measures. Rather this study aims to provide robust estimates for the likely recruitment and retention rates, and give an indication of the potential variability in the proposed outcome measures, which will in turn be used to inform the power calculation for a future definitive RCT. This is discussed further under statistical analyses.

### Recruitment and consent

#### Recruitment

General practices will be identified through contact details publicly available via practice websites and through contact lists provided by the health boards across Wales. Practices will be invited to participate using a standardised invitation via email addressed to the practice manager, along with information about the study. Practice managers will be asked to consult with their team and indicate their interest in participating in the study by responding to the email. They will also need to advise of their practice’s availability for potential workshop dates.

If no response to the initial email is received, a reminder email will be sent followed by a telephone call. A participant flow diagram can be seen in figure 2.

**Figure 2:** Participant flow diagram

Practices that take part in the study will be financially reimbursed for their time. We will establish contacts within regional primary care clusters, with health board staff and with R&D departments in order to maximise potential recruitment opportunities. The recruitment methods will be continually assessed and iteratively developed to determine the most appropriate recruitment strategy for a future definitive RCT.

#### Eligibility to participate

As feasibility is the main objective of this study, all types of general medical practice will be eligible for inclusion. This will aid intervention refinement and allow for a better understanding of what is feasible across a range of practices, and also why some practices may not be able to take part.

#### Inclusion

Any general practice in Wales is eligible for inclusion. The target audience for the intervention, based on a ‘whole team approach’, includes all practice staff members. These may include, for example, GPs, nurse practitioners, nurses, health care support workers (HCSWs), practice managers, administrators, receptionists or any other practice staff, clinical and non-clinical. We would aim to include any new forms of primary care organisations such as managed GP practice networks, or other general medical practice amalgamations, as well as traditional GP partnerships. However, some of these may be unstable practices with no regular GP staff, and as such, they may be difficult to recruit or find it difficult to participate. Practices participating in other research are also eligible for inclusion; they will be asked to notify us of any cancer-related studies they may be participating in via the baseline questionnaire.

#### Exclusion

There are no exclusion criteria.

#### Consent

Practice managers will be sent more detailed study information in the form of a ‘Research Information Sheet for Practices’ (RISP) and a link to an online baseline practice questionnaire after they have expressed an interest to take part in the study. Participating practice managers will need to indicate that they have read the study information and have agreed with consent statements on the first page of the electronic questionnaire before they can proceed. Prior to the commencement of the ThinkCancer! workshop, Participant Information Sheets (PIS) will be provided by members of the research team and written consent will be obtained from all participating members of staff. This will include consent to use anonymised data recorded on paper or audio-file during workshops and workshop feedback forms. At this time point, participants will also be given the option to provide their contact details should they be happy to be contacted for a telephone interview. Those who indicate that they would like to take part will be contacted at least two months after the intervention. Participants from practices randomised to the control arm, who do not take part in a workshop, will also have the opportunity to take part in a telephone interview and will be sent an invitation letter via their practice manager. Response to interview invitations and supplying of contact details will be taken as consent to be contacted and informed consent will be obtained verbally at the time of interview.

*NB Although we will initially contact practice managers, and they will most likely be the person who completes the questionnaire, this task may be delegated to another member of the practice team with a particular interest in the study*.

#### Pre-trial pilot

A local practice, known to the research team, has agreed to participate in a ThinkCancer! pilot workshop prior to full rollout of the feasibility study. The practice is an urban, large 12,000 patient training practice in a moderately deprived region of North East Wales. Data will not be collected or recorded for trial purposes and feedback from those participating will only be used to refine the intervention prior to its delivery across recruited practices.

### Randomisation and blinding

The general medical practice will be the unit of randomisation. Randomisation will be achieved online, through the remote randomisation centre at the North Wales Organisation for Randomisation Trials in Health (NWORTH) at Bangor University. The randomisation system will use a dynamic adaptive allocation algorithm^39^ to achieve randomisation. ThinkCancer! is an open trial where blinding of participants, researchers and the statistician is not possible due to the nature of the intervention and 2:1 ratio for randomisation.

#### Withdrawal criteria

Practices (and individuals within a practice) will be free to withdraw from the trial at any time, and their right to refuse participation will be respected throughout. We will seek to understand their reasons where possible. In terms of the primary outcome measures, as long as it is possible to collect the data, intention-to-treat analysis will be utilised, whether or not the intervention was received or adhered to.

### Data collection

The feasibility study will be used to rehearse data collection approaches and assess their ease of use. Data will be collected at time-points specific to each item and depending on the type of data. All data collected in this study will be anonymised.

#### Proposed clinical effectiveness outcomes

Data relating to the proposed primary outcomes for the future definitive RCT will be collected at baseline and 6 months after randomisation. Two week wait referral data and PCI data will be collected directly from participating practices via Case Report Forms containing full instructions on how to extract the data from practice IT systems. We will work with the Practice Manager, CSNC or other delegated individuals to achieve this. It is recognised that this is likely to be too short a follow-up period for meaningful differences to be observed, but the main purpose in this case will be to test the feasibility of collecting the data in this way. Additionally, we will explore the availability of 2WW data at health board level.

#### Practice Questionnaires

The baseline and endline practice questionnaires will be available online to both intervention and control practices and are to be filled out by the practice manager or other designated person, ideally in collaboration with the practice team; SurveyMonkey™ will be the most likely platform. The questionnaires will consist of closed questions and some open, free-text questions, and will be used to collect data for each individual practice on the practice characteristics and current systems, and existing practice systems relating to cancer diagnosis and safety netting. The baseline data may be used to inform some workshop planning - i.e. workshop content and delivery may be tailored to some extent to suit individual practice needs and circumstances. Further process evaluation data will also be collected from the practice questionnaires as they will incorporate questions exploring contextual factors known to influence the success of quality improvement approaches used to improve health care.^40^ The baseline questionnaire will be completed by all practices prior to randomisation. The endline questionnaire will assess any differences in practice, knowledge or systems in comparison with those measured at baseline and will be completed at 6 months post-randomisation.

Baseline measures will include the following:

▪ Demographic information and practice characteristics (practice size, research-accredited status, number of clinical and non-clinical staff members, whether a teaching practice, etc.)
▪ Practice culture (e.g. team structure, diversity of team member roles, team decision-making processes)
▪ Practice knowledge with regards to safety netting and cancer awareness
▪ Current safety netting systems in place, if any, including:
  ○ What systems are in place
  ○ How widely they are used within the practice
  ○ How safety netting issues are communicated:
    ▪ Between clinicians
    ▪ To the wider practice team
    ▪ To patients
  ○ How safety netting is recorded

#### Feasibility and piloting data

Recruitment, retention and questionnaire completion numbers will be recorded throughout the trial. Spreadsheet systems will be put in place to record practice responses and to track their progress in the trial (e.g. number of practices approached, whether they have responded to the initial invitation, whether they have agreed to be randomised, etc.). Separate spreadsheets will also record feasibility data relating to the workshop itself, such as participant numbers.

#### Post-workshop reflections and participant feedback

Data specific to the intervention will be collected via participant feedback and observation and reflections of the research staff. Participant feedback forms will be distributed to practice staff; these can be completed in paper format or online. Responses will be requested using a combination of Yes/No choices, Likert scales and free-text comments. The questions will cover a number of areas including acceptability, usefulness, learning outcomes and the potential to change practice.^41^

Relevant *ad hoc* communications with practices throughout the study will also be collected on a spreadsheet, which may contribute to understanding the intervention in terms of what works, why and how.

The same research team members will deliver the intervention in all practices; their observations will be collected and will inform any refinements of the intervention. Observations and reflections recorded by the research team may provide valuable data on the potential effects of contextual factors, site-by-site and component-by-component measures, and the appropriateness of individual questions included in the practice questionnaires. They will also describe the cancer safety netting plan proposed by the practice and whether the SSNAP tool is used.

The researchers will keep a diary, which will include a record of any modifications made to the intervention and data collection methods.

#### Health economics

Health economics data collection sheets will be completed by the researchers following each workshop, and costings specific to the practice will be recorded. We will also use the feedback forms to determine staff roles within the practice for costing purposes.

#### NoMAD instrument

At least two months after the intervention, participants who consented to be contacted will be sent a link to complete an adapted Normalisation Measure Development (NoMAD) instrument.^42^ This will assess the implementation of the cancer safety netting plan using Normalization Process Theory (NPT) principles, which may or may not include the SSNAP tool depending on uptake.

#### Telephone interviews

We will conduct qualitative telephone interviews lasting up to 30 minutes with a purposive sample of up to 45 clinical and non-clinical practice staff. Practice staff in both arms of the trial will be eligible to participate in the interviews as they will be invited to give feedback on all aspects of the trial process including the intervention where appropriate. The qualitative interviews are designed to achieve an in-depth understanding of the views and perceptions of practice staff involved in the trial. The interviews will allow participants to explain how they were able to utilise aspects of the trial and how they worked in practice. Informed consent will be obtained and interviews recorded and transcribed verbatim. The interviews will be semi-structured and will follow a pre-defined topic guide, although not every participant will be engaged with every section of the topic guide (i.e. only the specific areas of the topic guide that are relevant to an individual’s role and experience will be explored). Topics may include acceptability, safety netting, data collection, uptake of the intervention and SSNAP tool and implications. These interviews will occur at least two months after the intervention has taken place; control practices will be invited two months post-randomisation.

#### Adverse events

A risk assessment has found this trial to be low risk. Non-serious adverse events will not be collected. However, Serious Adverse Events (SAEs) will be reported that could be related to the intervention, as decided by the Chief Investigator (CI), and in line with current ICH-GCP Standard Operating Procedures.^43^ SAEs are defined as follows:

> “…an untoward occurrence that (a) results in death; (b) is life-threatening; (c) requires hospitalisation; (d) results in persistent or significant disability or incapacity; (e) consists of a congenital anomaly or birth defect; or (f) is otherwise considered medically significant by the investigator”^43^

We do not expect any related SAEs for this study.

### Data analysis

#### Quantitative analysis

A fully documented statistical analysis plan will be prepared by a registered trials unit and agreed by the co-investigators and approved by the trial governance structure, which will be known as the Trial Steering Committee (TSC).

Baseline characteristics will be summarised for all practices, the intervention group and control groups separately. Feasibility and process evaluation data such as practice recruitment rate, implementation and uptake of and adherence to the intervention, and follow-up rates will be summarised and presented as percentages.

Determining differences in clinical outcomes between the control and intervention is not the primary purpose of this study, therefore the focus of the results will be on the estimates of the treatment effects rather than statistical significance and as such, no hypothesis testing will be undertaken. As recommended in guidelines for good practice for the analysis of pilot studies,^32^ summary estimates of effects will be developed along with their 95% confidence intervals. Differences between the two comparison groups will be presented in the form of an unadjusted mean difference for continuous outcomes, and an odds ratio for binary outcomes. Exploratory analysis using ANCOVA for continuous outcomes and logistic regression for binary outcomes will consider adjustment for the stratification variables in assessment of the treatment effects.

Factors associated with the ability to implement the intervention will be tentatively explored using logistic regression with the focus on identifying deterministic barriers to implementation rather than probabilistic factors. The nature of the intervention may vary, directed by real-time feedback during the course of the trial and this will need to be taken into consideration during analysis.

As this is a feasibility study, there will be no imputation of missing data over and above any scoring rules established for the outcomes. This information will be used to feed into the suitability and applicability of the chosen outcome measures.

#### Economic Analysis

Alongside the statistical analysis plan, a Health Economics Analysis Plan (HEAP) will be produced setting out the objectives, methods and plans for dissemination of the health economics findings.^44^ The costing of the development of the ThinkCancer! Intervention will include researcher time, piloting, materials development, printing, publication, development of online materials, etc. Delivery costs of the ThinkCancer! Intervention will be determined based on the following:

▪ Online delivery format – live seminars/webinars, staff time and materials, exploration of whether health professional time should be collected in a full trial to reflect the co-production nature of CPD in own time or reflecting the opportunity cost of CPD in terms of time not spent on direct patient care activities.Mixed-format delivery – potential costs of a face-to-face/online delivery format across Wales in future after COVID-19.*Qualitative analysis*
▪ The transcribed telephone interviews, the free text responses from the feedback forms and the observational data, in text form, will be analysed for the process evaluation using Framework Analysis.^45^ Framework is a five-stage matrix-based system for analysing qualitative data which is highly appropriate for a feasibility study which is iterative in its development. Initially all transcripts and textual data will be read thoroughly by the same researcher who conducted the interviews to achieve data familiarisation and immersion. An index of emergent themes will then be created and data coded according to the index. Charts will be created according to the themes and coded data will be synthesised into the appropriate thematic charts. The completed charts will then be used for final stage which is in-depth interpretation.^45^

## TRIAL MANAGEMENT

The Sponsor is Bangor University. The study will be supported by the North Wales Organisation for Randomised Trials in Health (NWORTH), which is a fully registered Clinical Trials Unit.

There will be no on-site monitoring as there are no local research teams at sites. Therefore, the monitoring of data will have a more internal focus in the form of self-audits to ensure compliance with regulations.

### Trial governance

#### Operational group

The operational working group will be responsible for the overall conduct, supervision and progress of the study. They consist of the immediate research team, supported by a wider group of experts.

#### Trial Management group

The Trial Management group (TMG) will meet once a month, consisting of the operational group and a wider team of experts, including a PPI member. The group will be responsible for the overall management of the trial, and ensuring the study adheres to the protocol.

#### Trial Governance Structure

A Trial Steering Committee (TSC) committee will provide independent oversight for the study, ensuring it is conducted according to the standards set out by the HRA Research Governance Framework.^46^ As the study includes an element of ongoing intervention refinement, and is deemed low risk with very minimal likelihood of stopping early due to patient safety, a Data Monitoring and Ethics Committee (DMEC) will not be required.^47^

Meetings are expected to be biannual and the Sponsor and Funder will be updated following each meeting. The TSC will have an independent chairperson and at least three independent members including Patient and Public Involvement (PPI) representation, trial co-applicants, statisticians, health economist(s) and GPs.

### Data Management

A detailed data management plan will be written by NWORTH staff. This plan will include the definition of the data quality checks that will be performed on the data throughout the life course of the trial. These will include source data validation, random data checks and timelines for data entry.

#### Quality control

Quality control will be maintained through adherence to the study protocol, Betsi Cadwaladr University Health Board/Bangor University Standard Operating Procedures (SOPs), principles of Good Clinical Practice, research governance and clinical trial regulations.

#### Data protection and participant confidentiality

All investigators, trial site and research staff will comply with the requirements and regulations of the EU General Data Protection Regulation 2018 (GDPR) regarding the collection, storage, processing and disclosure of personal information and will uphold the Regulation’s core principles. All research staff involved will have up to date GCP training. Research data will be retained as per the Sponsor’s research data management policy. Bangor University is the data custodian.

#### Data archiving

As per the Sponsor’s research data management policy, research data and records will be archived along with the data management policy of the Sponsor.

In line with legal requirements, trial documents will be archived centrally at a secure facility with appropriate environmental controls and adequate protection from fire, flood and unauthorized access. Archived material will be stored in tamper-proof archive boxes that are clearly labelled. Electronic archiving will be provided by the Sponsor for post-project deposit and retention of data.

Destruction of essential documents will require authorisation from the Sponsor.

### Dissemination policy

On completion of the study a final report will be prepared for Cancer Research Wales.

Findings will be disseminated through various media, including open-access peer-reviewed publications, national and international conferences, the programme web pages, social media, and through an end-of-programme symposium for key stakeholders. Findings will also be disseminated to participating practice teams.

Publications arising directly from the WICKED programme and authorship on the final trial report will adhere to the BMJ guidelines on authorship and contribution, based on the International Committee of Medical Journal Editors Recommendations for the Conduct, Reporting, Editing, and Publication of Scholarly Work in Medical Journals (ICMJE) 2013.^48^

### Patient and public involvement

The study team recognises that the involvement of those with lived experiences will be vital in this research. Furthermore, a lay perspective is essential in the development and undertaking of research for the promotion of equality, diversity and transparency. Two patient representatives were initially recruited to the WICKED programme, one of whom has maintained active involvement in the study design, the development of the protocol and conduct throughout. Additionally, the trial PPI has been active in providing feedback on participant-facing documents. Two more PPI representatives have been recruited to the TSC through the North Wales Cancer Forum and have directly relevant experience. Their perspective as both a patient and a member of the public will inform the overall supervision of the trial.

## DISCUSSION

This study aims to test the feasibility of the ThinkCancer! intervention. The ThinkCancer! Study comprises Work Package 4 (WP4) of a programme of research called the Wales Interventions and Cancer Knowledge about Early Diagnosis (WICKED). The intervention will consist of a workshop aimed at the entire general practice team, as previous work packages have demonstrated the value of a whole-practice approach. If the intervention is shown to be feasible, we will proceed with designing a full-scale definitive trial.

This trial is especially relevant due to the current ongoing Covid-19 pandemic, which has had a major negative impact on suspected cancer referrals to secondary care, and will likely delay many cancer diagnoses and treatments, leading to poorer patient outcomes. One of the key strengths of this intervention is that it can be iteratively developed throughout the study period, which will ensure the future definitive trial will adopt an optimal approach. The design process of the study is also a strength in that a strong multidisciplinary team and advisory groups have been involved throughout. Furthermore, the mixed methods approach will allow us to capture a variety of data from this complex intervention. Furthermore, the planned rigour with which the study will be conducted is also a strength.

We are aware that some challenges in the recruitment lay ahead, but we plan to work with the various research infrastructures in Wales to overcome this. We plan to work closely with the Primary Care Specialty Lead and Primary Care Research Managers within Health and Care Research Wales, as well as with the individual health board R&D departments in order to maximise recruitment. We will also work closely with the practices that agree to take part in order to support their participation in the study.

Although safety-netting has garnered more attention in recent years, there currently are no recommendations on how best to do it.^26^ To our knowledge, there are no interventions targeting primary care with a focus on safety netting. In addition, involving the entire practice is a relatively novel approach, with great potential benefit. This study encompasses a multicomponent and complex behavioural change intervention comprising a multidimensional, interactive, tailored and whole-practice approach, which is timely and needed to optimise primary care approaches to the timely diagnosis of cancer.

## TRIAL STATUS

The trial is currently open for recruitment.

## Supporting information

Figure 1 - Logic Model

Figure 2 - Participant Flow Diagram

SPIRIT Checklist

## Data Availability

On completion of the trial, the final datasets generated and/or analysed will be available from the corresponding author on reasonable request. Access to the final datasets will be in accordance with governance policies, Good Clinical Practice (GCP) guidelines and funder arrangements.

## ABBREVIATIONS

2WW: Two Week Wait
AE: Adverse Event
CSNC: Cancer Safety Netting Champion
CSNP: Cancer Safety Netting Plan
DMEC: Data Monitoring and Ethics Committee
GCP: Good Clinical Practice
GDPR: General Data Protection Regulations
GP: General Practitioner
HCSW: Health Care Support Worker
HEAP: Health Economics Analysis Plan
HRA: Health Research Authority
MRC: Medical Research Council
NIHR: National Institute for Health Research
NWORTH: North Wales Organisation for Randomisation Trials in Health
PCI: Primary Care Interval
PCP: Primary Care Provider
PIS: Participant Information Sheet
PPI: Patient and Public Involvement
R&D: Research and Development
RCT: Randomised Controlled Trial
RISP: Research Information Sheet for Practices
REC: Research Ethics Committee
SAE: Serious Adverse Event
SOP: Standard Operating Procedures
TSC: Trial Steering Committee
WICKED: Wales Interventions and Cancer Knowledge about Early Diagnosis
WP: Work Package

## DECLARATIONS

### Ethics approval and consent to participate

This study has been approved by the Health Research Authority/Health and Care Research Wales (IRAS 256824) and Bangor University (School of Healthcare Sciences, 2019-16498). We did not seek REC Approval; this was not required as the study does not involve patients.

Participants are consented at two different time points in the study. Practice managers will consent to participate in the study on behalf of the general medical practice. If the practice has been randomised to the intervention, individual staff participants will be asked for their informed consent at the start of the workshop.

### Trial Sponsor

Bangor University. The Sponsor had no role in the design of this trial and will not have any role during its execution, analyses, data interpretation or decision to submit results.

Contact: Dr Huw Roberts. School of Healthcare Sciences, The Normal Site, Holyhead Road, Gwynedd, LL57 2AS, UK; Tel: 01248 383 136;

### Consent for publication

No participant identifiable information will be published.

### Indemnity

Bangor University currently has in place appropriate Clinical Trials Indemnity and Professional Indemnity insurance covering harm to participants arising from the management and/or the design of the research and member of the research team to conduct the research as per protocol. On sites that are not covered by the NHS Indemnity Scheme (e.g. GP surgeries in primary care), investigators/collaborators will need to ensure that their activity on the study is covered under their own professional indemnity. Health and Care Research Wales assisting with recruitment will have NHS contracts and will be responsible for ensuring they are appropriately insured.

### Authors’ contributions

RDN and CW are the Chief Investigators for the study. CW and RDN contributed to all aspects of this work including: conceiving and design of the study, obtaining initial and additional funding, intervention development, management of the study, obtaining approvals and writing the report.

SD is the Research Project Support Officer for the study. SD drafted the manuscript integrating comments from JR, AS, AH, NG, CW, RN, NW, RTE, BFA, RL, RJL, and ACS. SD was also involved in the development of the intervention and its adaptation in response to COVID-19.

AS was involved in the development of the intervention, its adaptation in response to COVID-19 and contributed to the protocol via the intervention description and by developing the logic model. AS also read and approved the final manuscript. The workshop materials were designed by AS, with assistance and advice from the wider ThinkCancer! team. AS is also the GP Educator and will be delivering the intervention.

JR is the trial manager for the study. She oversaw the writing of the protocol, facilitated gaining approvals from the seven health boards and contributed to adaptations of the methodology in response to COVID-19. JR also read and approved the final manuscript.

AH is the qualitative researcher for the study and contributed to the protocol via the embedded qualitative component and design of qualitative data collection materials. AH also contributed to gaining R&D approvals from the seven health boards. AH also read and approved the final manuscript.

RL was involved in the development of the intervention, the study design and the previous work packages leading up to the ThinkCancer trial. RL contributed to the methodological components of the protocol. RL read and approved the final manuscript.

NG is the Trial Statistician and contributed to the protocol via the statistical component. NG also read and approved the final manuscript. ZH is the Principal Statistician and will oversee all statistical analysis. ZH was involved in the design of the study and contributed to the protocol via the statistical component and the outcome measures. ZH read and approved the manuscript.

RTE is Lead Health Economist. BFA is Health Economics Research Support Project Officer and drafted the health economics analysis plan and is responsible for data analysis. RTE and BFA read and approved the manuscript.

RJL was involved in the intervention development and the previous work packages leading up to the ThinkCancer trial. RJL read and approved the final manuscript.

JH was involved in the intervention development and the previous work packages leading up to the ThinkCancer trial. JH read and approved the final manuscript.

ACS contributed to the design of the study and the development of the intervention. ACS also read and approved the final manuscript.

## Acknowledgements

The authors would like to thank Maggie Hendry for her significant contributions to the trial protocol on which this paper is based, as well as her contributions to the development of the intervention, and Stella Wright, for her contributions to the trial protocol and the participant-facing materials. We would also like to acknowledge Nic Nikolic for her work on the study costings. We would like to thank Jan Rose, Brian Lewin and Rosemary Birch, our patient representatives, for their valuable contributions to the development of the protocol and the participant-facing materials. We would also like to acknowledge the contributions from Dr Jane Heyhoe from the Yorkshire and Humber Patient Safety Translational Research Centre and Dr Caroline Green from RedWhale GPUpdate.

## ADDITIONAL MATERIALS

Additional file 1: WHO Trial Registration Data Set (Additional File 1 – WHO trial registration data.pdf)

Additional file 2: ThinkCancer! Logic Model (Additional File 2 – logic model.pdf)

Additional file 3: participant flow diagram (Additional File 3 – participant flow diagram.pdf)

Additional file 4: SPIRIT Checklist (Additional File 4 – SPIRIT.pdf)

