## Supplementary figures and images for "Protocol for a Feasibility study incorporating a Randomised Pilot Trial with an Embedded Process Evaluation and Feasibility Economic Analysis of ThinkCancer!: A primary care intervention to expedite cancer diagnosis in Wales"

### Figure 1 - Logic Model

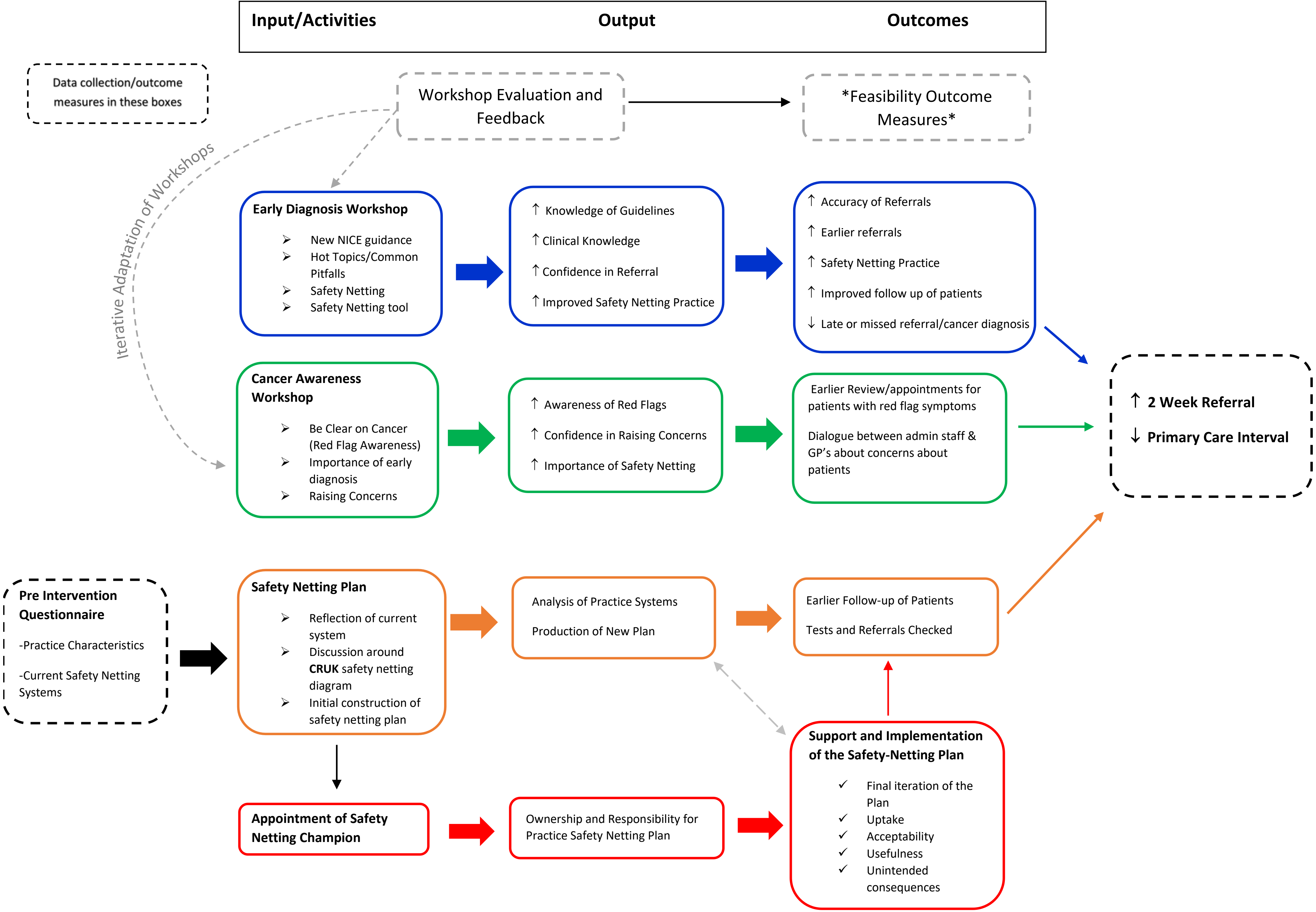

### Figure 2 - Participant Flow Diagram

Figure 2: Participant flow diagram

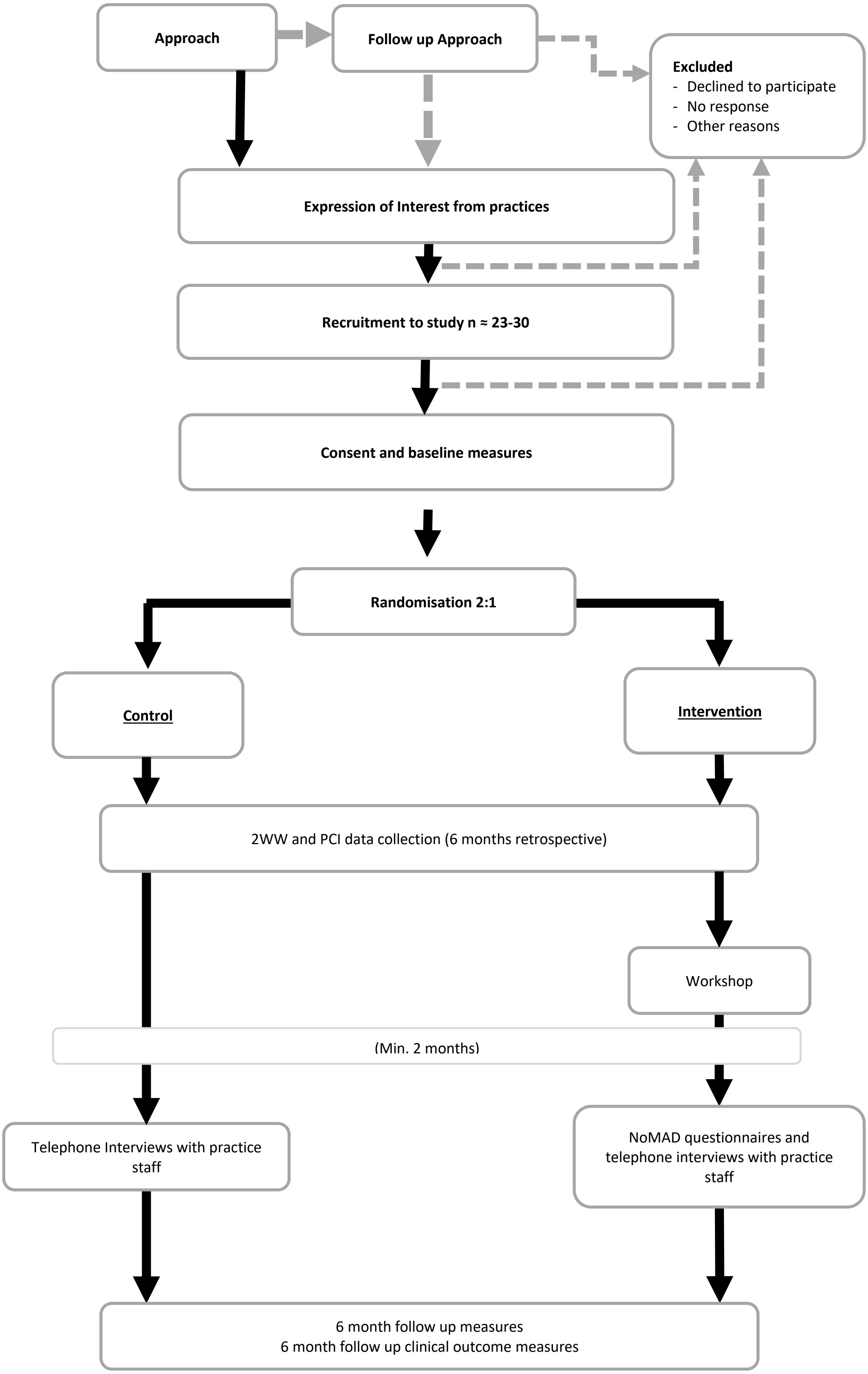
